# Anticipating racial/ethnic mortality displacement from COVID-19

**DOI:** 10.1101/2021.09.09.21263351

**Authors:** Stephen M. Kissler, Yonatan H. Grad

## Abstract

In 2020, life expectancy in the United States decreased by an estimated 1.5 years. Due to mortality displacement during the COVID-19 pandemic, life expectancy could soon rebound to above its pre-pandemic baseline. We estimated the size and duration of this anticipated rise in life expectancy through 2030. We found that this rebound could persist for years and will likely be most pronounced in minority populations who suffered the highest rates of mortality during the pandemic. Accounting for this artificial rebound will be critical to avoid funneling resources away from populations that still urgently need them.

## Main text

In 2020, life expectancy in the United States decreased by an estimated 1.5 years, mainly due to the COVID-19 pandemic.^1^ The decline was sharper for non-white Americans.^1^ Life expectancy is a key metric for determining overall well-being and disparities in risks borne by different populations. To place life expectancy statistics in the appropriate context in the coming years, we need to anticipate how they will vary due to the pandemic and account for mortality displacement.^2^

Mortality displacement occurs when a catastrophic event, such as a heat wave or an infectious disease outbreak,^2,6^ temporarily increases mortality rates in a population and is followed by a period of lower mortality rates. Such events usually pose the greatest risk to the most vulnerable members of the population. Mortality in these groups is thus accelerated by the event. After the event, the expected lifespan of the surviving population may increase, not because the well-being of any individual within the population has improved, but instead because the impact of vulnerable individuals who died prematurely on life expectancy calculations has been ‘displaced’ to the event. Life expectancy statistics therefore must be interpreted with caution.

COVID-19 results in higher mortality in those who might otherwise have had few life-years remaining—for example, those with obesity, diabetes, and heart disease, all of which are associated with lower life expectancy. Mortality displacement may thereby skew the surviving population toward those with longer remaining lifespans, resulting in a rebound in life expectancy even above the pre-pandemic baseline and in the erroneous conclusion that health and life expectancy have not only recovered but improved.

We estimated the size and duration of this artificial rise in life expectancy through 2030. Life tables from 2014-2018 and excess mortality statistics for 2020 were obtained from the National Center for Health Statistics, Centers for Disease Control and Prevention.^3,4^ Baseline lifespans were assigned to a theoretical cohort according to the expected number of remaining life-years by age and race/ethnicity in 2018, the most recent year with available data. Then, for each age and race/ethnicity group, excess mortality in 2020 was distributed among those with varying numbers of future life-years remaining according to a risk reduction constant *k*. Each additional future life-year at baseline came with a *k*-fold reduction in the probability of death in 2020, such that *k* = 1 yielded no difference in risk and *k* = 2 yielded a two-fold reduction (halving) of mortality risk for each additional future life-year. After removing the excess deaths, we followed the cohort forward through 2030, calculating life expectancy in each year. Full details are in the **Supplementary methods**. Code is available at https://github.com/gradlab/LifeExpectancy.

We found that any non-uniform distribution of excess mortality (*k* > 1) yielded an artificial increase in life expectancy after 2020 (**Figure 1**). Larger values of *k* yielded larger but shorter-lasting increases. For fixed values of *k*, the artificial increase was larger and more durable for Hispanics and non-Hispanic Blacks than for non-Hispanic Whites.

**Figure 1.**
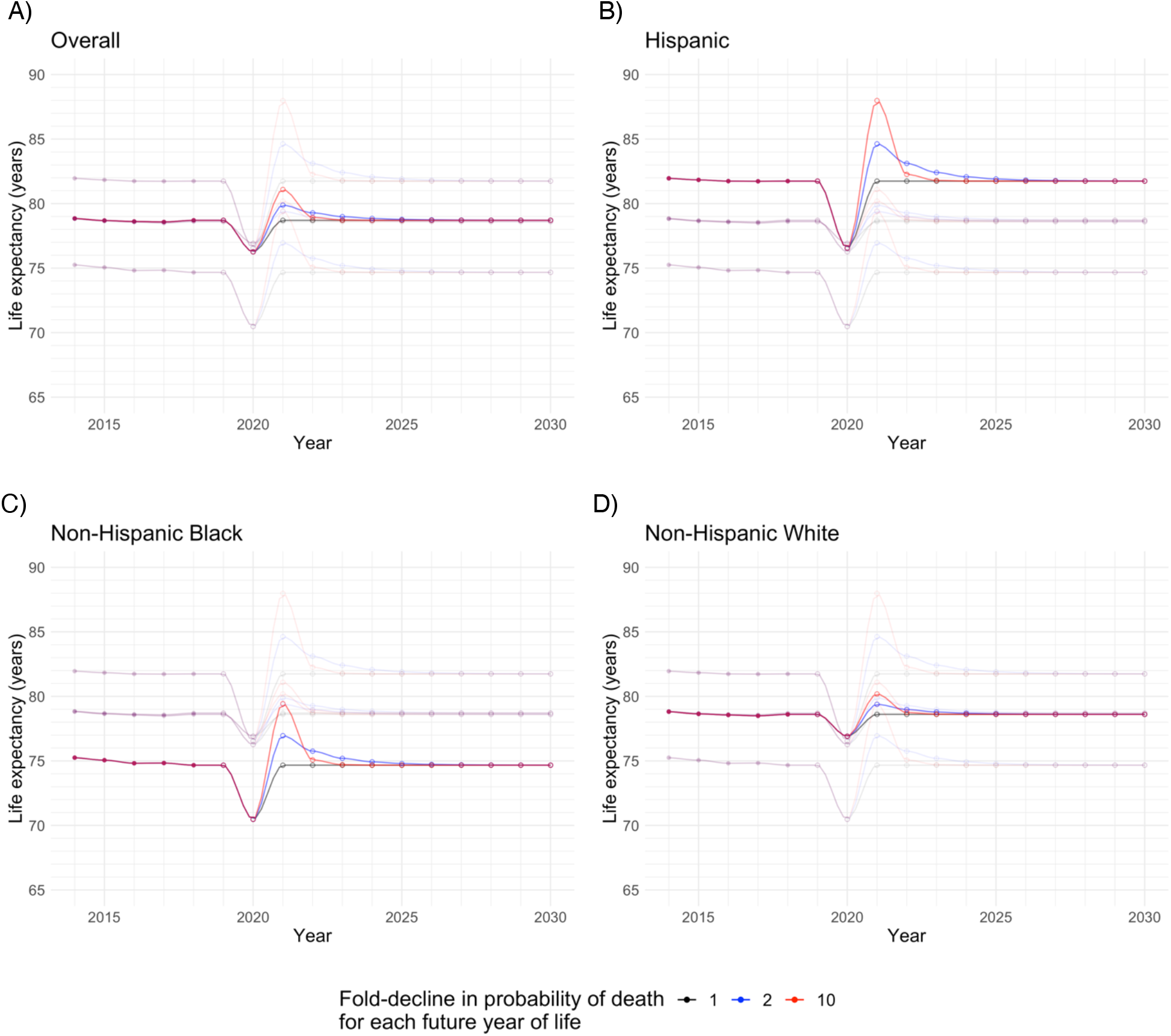
Decline and anticipated rebound in life expectancy due to excess mortality in 2020 by race/ethnicity. Solid lines depict the estimated life expectancy for (A) the US population overall, (B) the Hispanic population, (C) the non-Hispanic Black population, and (D) the non-Hispanic White population. Lighter lines depict the life expectancies for the other populations for reference. Colors represent a range of risk reduction coefficients *k*, where *k* = 1 represents no difference in 2020 mortality risk across expected future life-years, *k* = 2 represents a two-fold decline in 2020 mortality risk for each additional future expected life-year, and *k* = 10 represents a ten-fold decline in 2020 mortality risk for each additional future expected life-year. Solid points represent life expectancies calculated from existing life tables and open points represent projected life expectancies.

This artificial increase in life expectancy from mortality displacement could lead to incorrect claims about the extent to which health improved in the wake of the pandemic, especially for non-white populations. This could funnel resources away from populations that still urgently need them. The rebound may be counteracted by further declines in life expectancy due to continued transmission of SARS-CoV-2 and to long-term medical, social, and economic effects from the pandemic.^5^ Reporting multi-year life expectancies that incorporate the sharp decline in 2020 could give a more realistic view of collective well-being.

This study complements others that have documented declines in life expectancy due to the pandemic^5^ and that have attributed mortality displacement effects to other acute infectious diseases.^6^ The declines we estimated are somewhat larger than those reported in a provisional report by the Centers for Disease Control and Prevention^1^, possibly because those figures are adjusted for multiple biases in reported mortality for which data are not publicly available. Nevertheless, our figures are roughly in line with other estimates, and we anticipate that the overall patterns in future life expectancy will hold.

## Data Availability

Data and code are available at https://github.com/gradlab/LifeExpectancy

## Disclaimer

The findings, conclusions, and views expressed are those of the author(s) and do not necessarily represent the official position of the Centers for Disease Control and Prevention (CDC).

## Supplementary methods

Here, we describe the methods used to estimate the mortality displacement due to excess deaths in 2020. We begin with an overview of the four-step calculation and then provide an illustration.

### Mortality displacement calculation

#### 1) Calculate life expectancy for 2014 – 2019

Life tables for the United States for 2014–2018 were obtained from the National Center for Health Statistics (NCHS), Centers for Disease Control and Prevention (CDC).^3^ These tables report the probability of death between age *a* and *a + 1* conditional on survival to age *a*, denoted *q*_*a*_. To calculate life expectancy, we first calculated the unconditional probability of death between age *a* and *a + 1*, denoted *p*_*a*_, using the formula

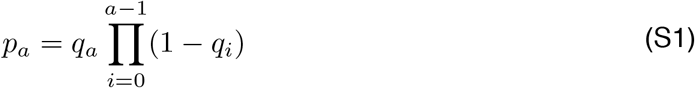

where *q*_*–1*_ is trivially defined to be 0 so that *p*_*0*_ *= q*_*0*_ (see “Illustration”, **Supplementary Table 1**). Then, the formula for life expectancy at birth *e*_*0*_ is

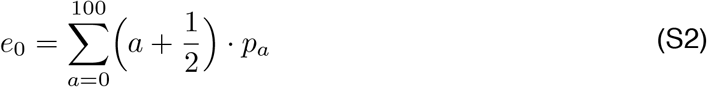

The additional 1/2-year within the parentheses in Eq. S2 reflects the assumption that the rate of death within an age-year is constant across that year.^3^ We calculated the overall life expectancy and the life expectancy for Hispanic, non-Hispanic Black, and non-Hispanic White individuals for 2014 – 2018 using the respective life tables for those demographics and years. Since data were not available for 2019, we used the corresponding life tables from 2018.

Note that the life tables are only defined through age 100, with a conditional probability of death of *q*_*100*_ = 1 at age 100. For the formal life expectancy calculations reported by the CDC, a logistic extrapolation procedure is used to extend the probability of death estimates through age 120, while for our calculations we retained the simplifying assumption that *q*_*100*_ = 1. This introduces a small discrepancy between our life expectancy calculations at the *e*_*0*_ values reported by the CDC, but the discrepancy is never greater than 2 months.

#### 2) Calculate the excess probability of death and life expectancy in 2020

The NCHS has also published estimates of excess deaths in 2020 relative to the 2015–2019 average.^4^ These are reported by racial/ethnic group and age groups (0–14, 15–19, 20–24, 25– 29, 30–34, 35–39, 40–44, 45–49, 50–54, 55–59, 60–64, 65–69, 70–74, 75–79, 80–84, 85+). We calculated fold-increase in the death rate at age *a* in 2020, *ξ*_*a*_, by dividing the deaths in 2020 by the mean annual deaths in 2015–2019 for the age group containing *a*.

Next, we calculated the probability of death at age *a* in 2020 conditional on survival to age *a, q*_*a*_*\**, using the formula

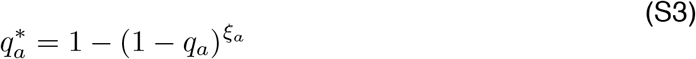

The excess probability of death in 2020 is *q*_*a*_*\* – q*_*a*_. The life expectancy in 2020 is obtained by calculating *p*_*a*_*\** from *q*_*a*_*\** using Eq. S1 and then substituting *p*_*a*_*\** for *p*_*a*_ in Eq. S2.

#### 3) Distribute excess deaths across future life-years

For each age *a*, we defined the distribution of future remaining life-years *n* at age *a* (*n* ≥ 0) in the absence of excess deaths, denoted *f*_*a*_(*n*), as

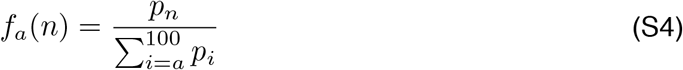

Note that *f*_*a*_(0) *= q*_*a*_, that is, the proportion of individuals with 0 remaining life years at age *a* is equal to the probability of death at age *a* conditional on survival to age *a*.

Next, we allocated the excess probability of death in 2020, *p*_*a*_*\* – p*_*a*_, among the future life-years *f*_*a*_(*n*) for *n* > 0 according to the scaling factor *k*, where *k* is the fold-reduction in risk of death in 2020 associated with each remaining future life-year. That is, a person of age *a* with *n* remaining life-years has a *k*-times greater risk of dying in 2020 than a person of the same age with *n+1* remaining life years. When *k* = 1, the risk of death in 2020 is constant regardless of remaining lifespan; in this case, there is no mortality displacement. When *k* > 1, there is mortality displacement since individuals with longer remaining lifespans are more likely to survive. Given *k*, the distribution of excess deaths must satisfy

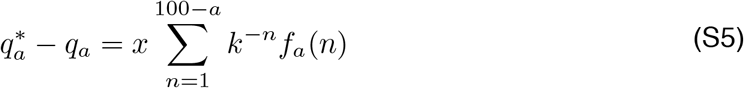

where *x* is a constant whose value is calculated by dividing *q*_*a*_*\* – q*_*a*_ by the sum on the right-hand side.

Finally, the proportion of individuals at age *a* with *n* life-years remaining (*n* > 0) after accounting for excess deaths in 2020 is

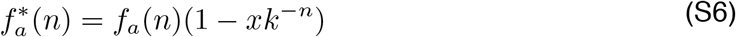

and these excess deaths are added to the proportion of individuals with no life-years remaining (*n* = 0), so that *f*_*a*_*(0) = *f*_*a*_*(0) + *q*_*a*_*\* – q*_*a*_.

#### 4) Calculate life expectancy for 2021– 2030

To project life expectancy in the years after 2020, we used the fact that *f\**_*a*_(*n*) in year *y* becomes *f\**_*a+1*_(*n*–1) in year *y+1*, with the appropriate normalization to account for deaths in year *y* so that

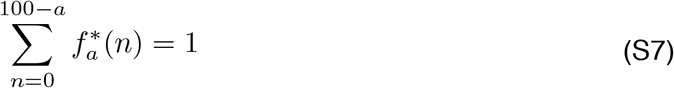

Calculating life expectancy in 2021 – 2030 then consists of three steps: (1) propagate surviving individuals to the next age *a* + 1, using the relation *f*_*a+1*_(*n*–1) in year *y* + 1 equals *f*_*a*_(*n*) in year *y*, and normalize the new *f*_*a*_(*n*) so that they sum to 1 according to Eq. S7; (2) calculate *p*_*a*_ using Eq. S1, noting that *q*_*a*_ = *f*_*a*_(0); and (3) calculate life expectancy according to Eq. S2.

### Illustration

We now provide an example of the above calculations. Code for this example is available at https://github.com/gradlab/LifeExpectancy. Consider an organism that lives to age 6 at most, where the probability of death at age 0 through age 6, conditional on survival, is *q*_*a*_ = {0.05, 0.1, 0.2, 0.3, 0.5, 0.7, 1}. **Supplementary Table 1** lists these conditional probabilities and the respective unconditional probabilities of death at each age, *p*_*a*_, calculated using Eq. S1. For this organism, the life expectancy (Eq. S2) is (0.5)(0.050) + (1.5)(0.095) + (2.5)(0.171) + (3.5)(0.205) + (4.5)(0.239) + (5.5)(0.168) + (6.5)(0.072) = 3.78.

**Supplementary Table 1.** Life table for a hypothetical organism that lives to age 6 at most. The conditional probability of death at age *a* (*q*_*a*_) is inferred from vital statistics; the unconditional probability of survival (*p*_*a*_) is calculated using Eq. S1; the fold-increase in death rate during year *y\** (*ξ*_*a*_) is inferred from mortality statistics; the conditional probability of death at age *a* in year *y\** (*q*_*a*_*) is calculated using Eq. S3; and the excess probability of death in year *y\** is *q*_*a*_* – *q*_*a*_.

| Age<br>($a$) | Probability of<br>death given sur-<br>vival to age $a$<br>($q_a$) | Probability<br>of survival<br>to age $a$<br>($p_a$) | Fold-increase in<br>rate of death<br>during year $y^*$<br>($\xi_a$) | Probability of<br>death given sur-<br>vival to age $a$ in<br>year $y^*$ ($q_a^*$) | Excess proba-<br>bility of death<br>in year $y^*$<br>($q_a^* - q_a$) |
| --- | --- | --- | --- | --- | --- |
| 0 | 0.05 | 0.050 | 1 | 0.050 | 0 |
| 1 | 0.1 | 0.095 | 1.05 | 0.104 | 0.004 |
| 2 | 0.2 | 0.171 | 1.1 | 0.217 | 0.017 |
| 3 | 0.3 | 0.205 | 1.2 | 0.348 | 0.048 |
| 4 | 0.5 | 0.239 | 1.3 | 0.593 | 0.093 |
| 5 | 0.7 | 0.168 | 1.4 | 0.814 | 0.114 |
| 6 | 1 | 0.072 | 1.6 | 1.000 | 0 |

Now, consider a catastrophic event that increases the rate of death in a single year *y\** by a factor of *ξ*_*a*_ = {1, 1.05, 1.1, 1.2, 1.3, 1.4, 1.6} for ages *a* in {0, 1, 2, …, 6}. The conditional probability of death at each age *a*, factoring in the catastrophe (*q*_*a*_*), is calculated using Eq. S3. The excess probability of death in year y* at each age a is *q*_*a*_* – *q*_*a*_ (**Supplementary Table 1**).

To keep track of the proportions of individuals with different numbers of remaining life-years, it is convenient to collect the *f*_*a*_(*n*) in an upper triangular matrix where *a* denotes the row and *n* denotes the column indexed from the diagonal. For the pre-catastrophe period, this matrix is

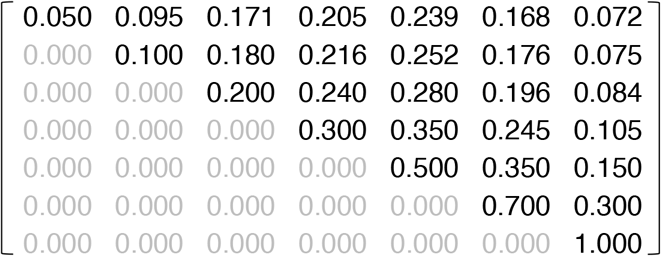

The entry in row *a* and column *j* (indexed from the diagonal, where *j* = 0 along the diagonal) represents the proportion of individuals of age *a* with *j* remaining years left to live. Note that the diagonal entries *f*_*a*_(0) equal the conditional probabilities of death *q*_*a*_ and the first row equals the unconditional probabilities of death *p*_*a*_ (**Supplementary Table 1**). Note also that each row sums to 1. These values are calculated using Eq. S4. We will refer to this object as the “life matrix”.

Next, we distribute the excess probability of death, *q*_*a*_*– *q*_*a*_, across row *a*. This only applies to the off-diagonal elements (*n* > 0); trivially, the individuals who would have died anyway in year *y\** still die (denoted by the diagonal elements), and so excess death must be taken from the individuals who otherwise would not have died in year *y\** (the off-diagonal elements). Consider the situation where *k* = 2, that is, the probability of death halves with each remaining future life-year. Our first task is to calculate the scaling factor *x* (Eq. S5). For this illustration, there is no difference in risk of death in year *y\** for individuals aged *a* = 0 (*ξ*_*0*_ = 1, *q*_*0*_*\** – *q*_*0*_ = 0), so we begin with the second row (*a = 1*). Here, the excess 0.004 probability of death must be distributed across the off-diagonal elements. We calculate *x* using Eq. S5:

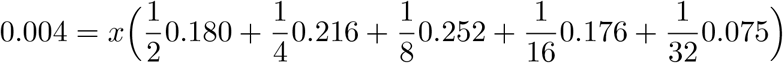

Solving for *x* gives *x =* 0.021 (the actual value is x = 0.025 if all the above calculations are done to machine precision instead of rounded). Then, using Eq. S6, we calculate the remaining proportion of individuals of age *a* with *n* remaining life-years after adjusting for excess death. For example, the remaining proportion of individuals of age *a* = 1 with one remaining life-year (*f*_*1*_*(1)) is *f*_*1*_*(1) [1 – x/2] = 0.180 [1 – 0.025/2] = 0.178. Repeating this calculation for all off-diagonal entries, and adding the excess deaths to the diagonal entry, yields the following life matrix that accounts for excess deaths in year *y\**:

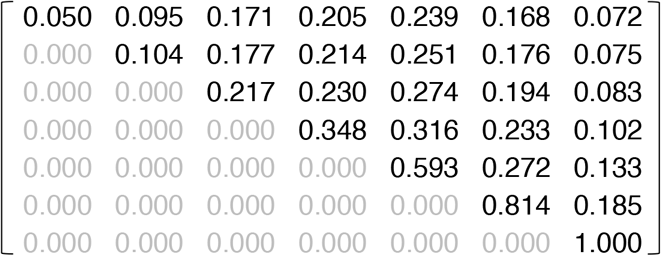

Note that the diagonal entries of this new life matrix equal the conditional probabilities of death in year *y*, q*_*a*_*\** (**Supplementary Table 1**), allowing us to calculate the life expectancy in year *y\** using Eq. S1 and S2.

Finally, we propagate surviving individuals forward. In each year, the individuals with zero remaining life-years die, leaving those with one or more remaining life-years to progress to the next age *a*. This is accomplished by setting the diagonal entries in the life matrix to 0 (death), shifting the rows down by one (aging), and re-normalizing the rows so that each row sums to 1. We fill in the first row of the life matrix using the baseline unconditional probabilities of death, *p*_*a*_. For year *y\** + 1, this yields the following life matrix:

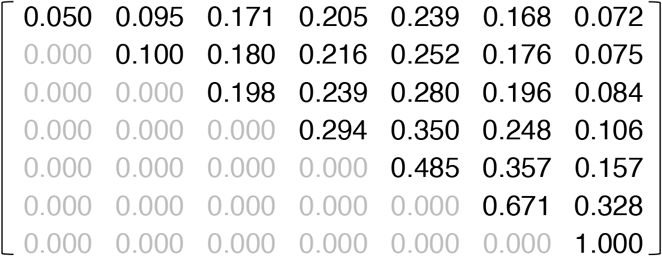

Again, the new life expectancy is calculated using the diagonal elements of this matrix, which represent *q*_*a*_ for the year following the catastrophe, and applying equations S1 and S2. This propagation may be repeated for an arbitrary number of years. The life expectancy for this organism in years *y\** – 1 through *y\** + 4 with *k* = 2 is {3.779 3.608 3.804 3.783 3.780 3.779}, showing an effect from mortality displacement where life expectancy declines in year *y\**, rebounds in year *y\** + 1, and finally settles back to roughly its baseline value in year *y\** + 4.

